# Evaluation of Semaphorin 3G (SEMA3G) as a biofluid biomarker of cerebral amyloid angiopathy: evidence from neurodegenerative disease and memory clinic cohorts

**DOI:** 10.64898/2026.11.18.26368450

**Authors:** Johannes C. Michaelian, Lisa Shi, Michael Barnett, Dongang Wang, Chenyu Wang, Sharon L. Naismith, Eleanor Drummond

**Affiliations:** Healthy Brain Ageing Program, Brain and Mind Centre, School of Psychology, Faculty of Science, The University of Sydney, Camperdown, NSW, 2050, Australia; Charles Perkins Centre, The University of Sydney, Camperdown, NSW, 2050, Australia; School of Medical Sciences, The University of Sydney, Camperdown, NSW, 2050, Australia; Brain and Mind Centre, The University of Sydney, Camperdown, NSW, 2050, Australia; Sydney Neuroimaging Analysis Centre, Camperdown, NSW, 2050, Australia; Sydney Medical School, Faculty of Medicine and Health, The University of Sydney, Camperdown, NSW, 2050, Australia

**Keywords:** Cerebral amyloid angiopathy (CAA), biomarker, plasma, cerebrospinal fluid, Semaphorin-3G (SEMA3G), Alzheimer’s disease

## Abstract

**Introduction:** Cerebral amyloid angiopathy (CAA) is characterised by amyloid-beta (Aβ) deposition in cerebral vessels, and frequently co-occurs with Alzheimer’s disease (AD). Diagnosis based on MRI-based Boston criteria is not definitive, and CAA biofluid biomarkers are needed. Semaphorin 3G (SEMA3G) was recently shown to be selectively enriched in CAA-affected vessels, and correlated with CAA severity. We therefore evaluated SEMA3G as a biofluid biomarker of CAA.

**Methods:** SEMA3G profiles were examined across diagnoses in the Global Neurodegeneration Proteomics Consortium dataset (GNPC) in cerebrospinal fluid (CSF) (N=1982) and plasma (N=6917). SEMA3G was then evaluated in the Healthy Brain Ageing (HBA) memory clinic cohort (N=161) with 3T MRI, where lobar cerebral microbleed burden was used as a proxy for CAA likelihood, with participants grouped by microbleed count (0, 1-2, or ≥3).

**Results:** CSF and plasma SEMA3G were significantly lower in AD and frontotemporal dementia (FTD) compared to cognitively unimpaired controls in GNPC, and were associated with hypertension. Longitudinally, plasma SEMA3G declined significantly over time in mild cognitive impairment (MCI) and AD. In the HBA cohort, plasma SEMA3G did not differ by cerebral microbleed burden on MRI, nor correlate with plasma Aβ isoforms and white matter hyperintensities.

**Discussion:** Although CSF and plasma SEMA3G levels were significantly lower in AD and FTD, plasma SEMA3G was not associated with CAA-associated MRI features in individuals at risk of AD. While these findings do not support plasma SEMA3G for identifying individuals with higher CAA-associated burden, its potential utility warrants further evaluation in genetically susceptible or neuropathologically confirmed CAA cohorts.

**Research-in-Context:**

1. **Systematic review:** We reviewed the literature using traditional sources (e.g. PubMed). SEMA3G has been shown to be selectively enriched in CAA-affected blood vessels across multiple brain regions and neurodegenerative conditions, but not in amyloid plaques or in CAA-negative blood vessels, highlighting its specificity for CAA. To date, no studies have examined whether SEMA3G is a viable central or peripheral biofluid biomarker of CAA.
2. **Interpretation:** CSF and plasma SEMA3G were consistently lower in AD and FTD, and plasma SEMA3G declined significantly over time in MCI and AD. However, plasma SEMA3G was not associated with features of CAA (lobar cerebral microbleeds, white matter hyperintensities, Aβ40), suggesting that plasma SEMA3G has limited utility in identifying individuals with higher CAA-associated burden or elevated ARIA risk.
3. **Future Directions:** Future studies should evaluate SEMA3G’s potential as a biomarker of CAA in genetically-susceptible or neuropathologically confirmed CAA cohorts.

**Highlights:**

- In the GNPC cohort, CSF and plasma SEMA3G were significantly lower in AD and FTD at baseline, and plasma SEMA3G decreased significantly over time in MCI and AD.
- A cluster of AD cases had particularly high levels of plasma SEMA3G that exceeded all other groups, but the biological cause of this remains unknown.
- In a memory clinic cohort, plasma SEMA3G did not differ significantly between participants with no (0), low (1-2), or higher (≥3) cerebral microbleed burden on MRI or correlate with other neuroimaging markers (i.e., white matter hyperintensities) of CAA.
- Plasma SEMA3G levels were inversely correlated with plasma NfL in the higher (≥3) cerebral microbleed burden group.

## 1. Background

Cerebral amyloid angiopathy (CAA) is a common age-related vascular disease characterised by the deposition of amyloid-beta (Aβ) within cortical and leptomeningeal blood vessels [1]. CAA frequently co-exists with Alzheimer’s disease (AD) pathology [2], and is a major contributor to lobar intracerebral haemorrhage, cerebral microbleeds and cortical superficial siderosis [1]. CAA can cause cognitive impairment and brain injury independently from AD [3], and can co-exist with other age-related neurodegenerative conditions, such as Parkinson’s disease (PD), although it is less common [4, 5]. In light of recently approved amyloid-lowering therapies for AD (i.e., lecanemab and donanemab), CAA has emerged as a key therapeutic constraint, given its strong association with an increased risk of amyloid-related imaging abnormalities (ARIA), with important implications for treatment eligibility, monitoring, and patient safety [6, 7].

While a definitive diagnosis of CAA requires post-mortem neuropathological examination, *in vivo* classification currently relies on the Boston criteria, which incorporate characteristic magnetic resonance imaging (MRI) features, including strictly lobar haemorrhagic lesions (i.e., intracerebral haemorrhage, cerebral microbleeds and cortical superficial siderosis), and white-matter features (i.e., multispot white matter hyperintensities) to classify either possible or probable CAA [8, 9]. Although widely adopted, these criteria are limited by specificity, specialised imaging and expert interpretation, hindering scalability and longitudinal assessment and monitoring [8]. These limitations underscore the need for more accessible, cost-effective and scalable approaches to characterising CAA and, importantly, for sensitive and specific biomarkers capable of detecting underlying cerebrovascular amyloid pathology.

Our group has recently shown Semaphorin 3G (SEMA3G) to be selectively and significantly enriched in CAA-affected blood vessels [10, 11]. Notably, SEMA3G was not enriched in amyloid plaques or in CAA-negative blood vessels in the same cases, highlighting its specificity for CAA-positive vessels. In a follow-up study, SEMA3G was significantly enriched in CAA-positive blood vessels in the inferior temporal cortex, primary visual cortex, and superior frontal cortex in both the parenchyma and leptomeninges [10]. In addition, the specificity of SEMA3G for CAA-positive vessels was observed across AD, Pick’s Disease, Progressive Supranuclear Palsy, Corticobasal Degeneration, and Lewy Body Disease, regardless of co-pathology. Preferential binding of SEMA3G with Aβ40 causing delayed fibril formation was also confirmed *in vitro* [10]. SEMA3G, a member of the class 3 Semaphorin family, plays a role in endothelial signalling, vascular patterning, and neurovascular unit integrity [12]. Its selective enrichment in amyloid-laden cerebral vessels therefore classifies it as a tissue biomarker of CAA, and suggests a close link to CAA-related vascular pathology [11]. This raises the possibility that SEMA3G may be detectable as a selective biofluid biomarker of CAA-related vascular pathology. To date, no studies have evaluated CSF or plasma SEMA3G as a potential biomarker of CAA.

In this study, we aimed to determine if SEMA3G is significantly altered in CSF and plasma across neurodegenerative conditions, including within and between groups likely to be at increased risk of CAA. To achieve this, we adopted a dual cohort approach, leveraging the recently published Global Neurodegeneration Proteomics Consortium (GNPC) dataset [13], comprising 18,645 participants across 23 cohorts, alongside the Healthy Brain Ageing (HBA) memory clinic cohort, a deeply phenotyped population. The GNPC cohort enabled large-scale, harmonised evaluation of CSF and plasma SEMA3G levels across neurodegenerative conditions, while the HBA cohort provided detailed whole brain 3T MRI assessment of CAA-associated features (i.e. lobar cerebral microbleed burden and white matter hyperintensities), comprehensive medical assessment, and blood-based biomarker profiling to characterise underlying neuropathological processes. Given the high occurrence of CAA in AD [2, 14], we hypothesised that SEMA3G would be significantly altered in the CSF and plasma of a subset of AD cases, reflecting those with CAA. We further hypothesised that SEMA3G would be associated with Aβ biomarkers and cardiovascular comorbidities.

## 2. Methods

### 2.1 The Global Neurodegeneration Proteomics Consortium

The Global Neurodegeneration Proteomics Consortium is the largest harmonised SomaScan 7k proteomics dataset of individuals from 23 cohorts with neurodegenerative diseases across the USA, UK and Europe. Proteomics data from each site was processed separately, where between-sample normalisation was performed using median signal intensities. V1 of the dataset included 53 clinical variables covering demographic, diagnostic, cognitive and comorbidity data [13]. Current (active) or a past history of comorbidities relevant to the participant at the time of sample collection were recorded. Comorbidities covered alcohol use, smoking/tobacco use, traumatic brain injury, cancer, chronic obstructive pulmonary disease, and a range of cardiovascular (stroke, transient ischaemic attack, congestive heart failure, myocardial infarction, atrial fibrillation, angina and hypertension), metabolic (diabetes and hyperlipidaemia) and mood (depression and anxiety) disorders.

Site-provided diagnoses for AD, frontotemporal dementia (FTD), PD and amyotrophic lateral sclerosis (ALS), as well as information on recruited controls and mild cognitive impairment (MCI) were retained. As specific diagnostic methods vary across the sites, a harmonised clinical diagnosis variable was provided by GNPC that was based on the Clinical Dementia Rating (CDR), and Mini-Mental State Examination (MMSE) or Montreal Cognitive Assessment (MoCA) scores for those with no formal diagnosis or CDR information: cognitively normal controls (CDR = 0 or confirmed recruited control participant), MCI (CDR = 0.5 or a confirmed diagnosis), Dementia (CDR ≥ 1 or a confirmed diagnosis of AD or FTD), and other neurodegenerative conditions (confirmed PD or ALS diagnosis).

Cases were filtered based on the following inclusion criteria: diagnosis with one unique neurodegenerative disease (either AD, FTD, PD or ALS) or MCI. Concurrent comorbidities were acceptable. Those with multiple neurodegenerative conditions were excluded. For plasma samples, controls were limited to those defined as a recruited control and confirmed absence of all other neurodegenerative diseases upon screening. As there were no recruited controls for cerebrospinal fluid (CSF), CSF control samples were defined by the absence of any neurodegenerative disease or MCI and a CDR of 0. All CSF and ethylenediaminetetraacetic acid (EDTA) plasma that passed quality control (based on the “quality_control” column) at baseline (earliest time point, t0) were selected. To ensure single sampling, only distinct sample and patient IDs were retained (based on “sample_id” and “person_id” columns, respectively). SOMAmer SL012850 and Sequence ID seq_5628_21 were used to identify SEMA3G. Only participants with complete SEMA3G, age, sex and BMI (plasma only) were retained for cross-sectional analyses.

To evaluate plasma SEMA3G trends over time across different diagnoses, a longitudinal GNPC dataset was created, filtering for cases with complete age and sex data, and a diagnosis with one unique neurodegenerative disease (either AD, FTD, PD or ALS) or MCI. As controls were only available at baseline, they were not included in this longitudinal cohort, and either MCI or AD was made the reference group depending on the model. The cohort was further filtered for cases with at least two timepoints (based on the “person_id” and “visit” columns), and who passed quality control (based on the “quality_control” column). As the duration between each visit was inconsistent between and within participants, a time (years) since baseline variable for each timepoint was calculated based on the participant’s age (age_at_visit), and used to control for mixed effects models. Age was measured at each visit, and some participants progressed from MCI to AD.

### 2.2 Healthy Brain Ageing clinic participants and case selection

Participants were recruited from the Healthy Brain Ageing (HBA) clinic, a specialist memory clinic in Sydney, Australia. Requiring a GP or medical specialist referral, the clinic provides comprehensive neuropsychological and medical assessments for older adults aged ≥50 years. Exclusion criteria for participation at the HBA clinic include head injury with loss of consciousness > 30 min, neurological disease (e.g., Parkinson’s disease) or non-affective psychiatric illness (e.g., schizophrenia), intellectual disability, insufficient English for neuropsychological assessment, current or previous substance misuse that may interfere with assessment, or a previously established diagnosis of dementia and/or MMSE < 20. All participants provided written consent prior to study participation, and all research activities were approved by the University of Sydney Human Research Ethics Committee (Protocol numbers: 2012/1873 and 2019/HE000271). All study methods were conducted in compliance with the Declaration of Helsinki.

Case selection for the present study was based on data availability and analytical feasibility. From the broader Healthy Brain Ageing (HBA) clinic cohort, participants were included if they had complete clinical characterisation, available blood samples, and high-quality neuroimaging data relevant to the study aims. Within this eligible subset, cases were further selected based on the availability of stored plasma suitable for downstream biomarker analyses.

### 2.3 Blood collection for HBA cohort

Fasting venous blood was collected on the morning (between 8:30 and 9:15 AM) of clinical assessment into ethylenediaminetetraacetic acid (EDTA) vacuum tubes. Within 30 minutes of collection, samples were centrifuged at 1,800 × g for 10 minutes at 4 °C. Plasma was aliquoted into cryovials and immediately stored at-80 °C until analysis.

### 2.4 Biomarkers for HBA cohort

All samples underwent two freeze-thaw cycles in total at the time of analysis. Plasma SEMA3G levels were measured in duplicate with a commercially available ELISA kit (MyBioSource, #MBS164724) in undiluted plasma according to the manufacturer’s instructions. All analyses were performed blinded to the clinical diagnosis. The intra-assay variation was <5%. Measurements of samples with a CV above 20% were repeated. The inter-assay CV, as indicated by an internal standard sample run on every plate, was <20%. Plasma concentrations of pTau217, neurofilament light chain (NfL), glial fibrillary acidic protein (GFAP), Aβ40, and Aβ42 were quantified on the Quanterix Simoa HD-X platform (Quanterix, Billerica, MA, USA). Plasma pTau217 concentration was measured using the Simoa ALZPath pTau217 kit (lot #504400), whereas plasma NfL, GFAP, Aβ40, and Aβ42 concentrations were measured in two analytical batches using the Simoa Neurology 4-Plex E (N4PE) assay kit (batch 1, 503819; batch 2, 504138). Across all kits and batches, samples were run according to the manufacturer’s instructions using the 4x dilution protocol, with values automatically adjusted for dilution by the instrument. All samples were assayed in duplicate; if the coefficient of variation (CV) exceeded 20%, measurements were repeated in duplicate, and the final value was derived from 3 of 4 replicates. To account for lot-to-lot variation between Simoa Neurology 4-Plex E (N4PE) assay kits across batches 1 and 2, 48 randomly selected samples from batch 1 were re-assayed using the newer Simoa Neuro 4-Plex E kit (lot number 504138) as bridging samples. Linear regression models were then fitted for each analyte (batch 1 vs batch 2 values) and the coefficients were applied to re-scale all batch 1 values to the batch 2 reference scale, generating a harmonised dataset of NfL, GFAP, Aβ40 and Aβ42 values.

Relevant to this study, participants were classified as pTau217-negative (plasma pTau217 concentration <0.4 pg/mL) or pTau217-positive (plasma pTau217 concentration ≥0.64 pg/mL) based on reference ranges providing 95% sensitivity and specificity in detecting amyloid positron emission tomography (PET) positivity [15]. Participants with pTau217 concentrations in between the cut-off ranges (0.4-0.64 pg/mL) were considered “indeterminate”.

### 2.5 Apolipoprotein E genotyping for HBA cohort

*APOE* genotyping was performed using DNA extracted from peripheral blood mononuclear cells. Genotyping was conducted at the Ramaciotti Centre for Genomics, UNSW Australia (NATA ISO/IEC Accreditation Number #20417) using the UK Biobank Axiom^TM^ Array, which probes single nucleotide polymorphisms (SNPs) rs429358 and rs7412 in the *APOE* gene. Where either SNP returned a no call (n = 1), genotype was imputed from proxy SNPs on the same array in strong linkage disequilibrium (LD) with the target SNP, with LD estimates taken from LDlink (1000 Genomes Phase 3, European population). rs7412 was imputed from rs1065853, and rs429358 from a combination of rs4420638, rs769449 and rs6857. Genotyping was performed blinded, and participants were classified as *APOE* ε4 carriers (≥1 ε4 allele) or non-carriers.

### 2.6 Neuroimaging

All participants underwent MRI on a 3T GE Discovery MR750 scanner (GE Medical Systems, USA). An 8-channel phased-array head coil was used before July 2019 and a 32-channel head coil thereafter. The protocol included a sagittal 3D T1-weighted BRAVO spoiled gradient-recalled echo sequence (before/after July 2019: TR/TE = 7.2/2.8 vs 7.4/3.0 ms; flip angle 12° vs 11°; 0.9 vs 1.0 mm isotropic voxels) and a sagittal 3D FLAIR sequence (TR/TE = 8000/160 vs 7002/159 ms; 1.2 vs 1.0 mm slices). Susceptibility-weighted imaging was acquired with an axial 3D multi-echo T2*-weighted gradient-echo sequence (SWAN; TR/TE = 37.1/23.1 ms; flip angle 15°; acquisition matrix 320 × 224; FOV 224 mm; slice thickness, 2 mm).

All image analyses were performed at the Sydney Neuroimaging Analysis Centre (SNAC; Sydney, Australia). White matter hyperintensities (WMH) were segmented on 3D FLAIR images using iQ-solutions™ (iQ-COG™, version 2.0.1), a fully automated platform cleared by the FDA (510(k) K231929) [16]. Cerebral microbleeds (CMB) were detected on 3D susceptibility weighted images (SWI) using a customised in-house research U-Net model built on the same framework. All automated outputs underwent quality assurance by SNAC analysts. As this was the first study to utilise cerebral microbleed counts generated by the automated algorithm, all scans were also manually assessed independently by a single neurologist with expertise in cerebral microbleed identification and characterisation. The automated algorithm identified more lobar microbleeds than manual assessment (313 vs 156), reflecting a higher number of detected lesions, and yielded a higher mean count per participant (2.09 [SD 11.32] vs 1.04 [SD 5.45], *p* < 0.001); nevertheless, counts derived using the two methods were strongly correlated (Pearson’s r = 0.986, *p* < 0.001).

### 2.7 Statistical analysis

#### 2.7.1 GNPC

BMI ranges provided were converted into underweight (< 18.5), normal (18.5 – 24.9), overweight (25.0 – 29.9) and obese (30.0 and above) categories. Participants with missing age, sex and SEMA3G were excluded to avoid imputation. Univariate ANOVA with Tukey’s post hoc or chi-squared with pairwise proportion tests were used to determine group differences in cohort characteristics. ANCOVAs adjusting for age, sex, and categorical BMI (plasma only) were performed in subsequent models. Tukey’s post hoc was used to determine significant pairs. All model assumptions were checked, and normality was ensured using transformations from the bestNormalize package in R. Mixed effects models were used to assess how plasma SEMA3G tracked over time across the various diagnoses, adjusting for age and sex only due to high missingness in BMI, and sample and years since baseline as random effects (sema3g ∼ years_since_baseline * group + age_at_visit + sex + bmi_categ + (1 + years_since_baseline | person_id)). All assumptions were checked, and a spline term for time supported linear approximation of change over time. Rank-based regression adjusting for the same covariates was used to assess associations between SEMA3G, all proteins and comorbidities. Bonferroni false discovery rate (FDR) correction was used where appropriate. Kruskal-Wallis tests were used to determine *APOE* group differences in SEMA3G, with Dunn’s multiple comparisons adjusted with Benjamini-Hochberg method. Analyses were conducted in R (version 4.5.2) and RStudio (version 2025.09.2+418) on AD Workbench.

#### 2.7.2 HBA

HBA participant characteristics were summarised as mean (standard deviation) for continuous variables and frequency (percentage) for categorical variables. Differences across MRI-derived CAA-burden groups, defined by 0 lobar cerebral microbleeds (low MRI evidence of CAA), 1-2 lobar cerebral microbleeds (increased likelihood of CAA) or ≥ 3 lobar cerebral microbleeds (higher CAA-associated haemorrhagic burden and elevated ARIA risk), were assessed using one-way analysis of variance, Kruskal-Wallis tests, or chi-squared tests, as appropriate, with significant omnibus tests (i.e., one-way ANOVA) followed by pairwise t-tests or Dunn’s tests. Spearman’s rank correlations using pairwise-complete observations examined associations between plasma SEMA3G, other plasma biomarkers, and neuroimaging measures in the overall sample and within each microbleed-burden group. All tests were two-sided, with *p* < 0.05 considered statistically significant. Analyses and visualisations were performed using R in RStudio (version 2026.08.1+195.pro1).

## 3. Results

### 3.1 GNPC cohort characteristics

A total of 1982 cases were included in the CSF analysis (495 control, 838 MCI, 283 AD, 85 FTD, 275 PD and 6 ALS) and 6917 cases were included in the plasma analysis (1211 control, 2113 MCI, 2653 AD, 180 FTD, 516 PD, 244 ALS) for which SEMA3G levels were quantified via SomaScan. CSF and plasma cohort characteristics, as well as median SEMA3G, NfL and GFAP are shown in Tables 1 & 2. The CSF cohort was on average 66.5 years of age, with 57.9% being female, of normal BMI, 10.9 years of education, with average scores of 24.8 and 23.5 on MMSE and MoCA, respectively, and 34.3% carrying at least one *APOE* ε4 allele. The plasma cohort was on average slightly older at 71.4 years of age, 54.5% female, of normal BMI, 12.6 years of education, with average scores of 25.3 and 18.7 on MMSE and MoCA, respectively, and 44.8% carrying at least one *APOE* ε4 allele.

**Table 1.** GNPC CSF cohort characteristics with biomarker levels.

|  | **Total sample (N = 1982)** | **Control (n = 495)** | **MCI (n = 838)** | **AD (n = 283)** | **FTD (n = 85)** | **PD (n = 275)** | **ALS (n = 6)** |
|---|---|---|---|---|---|---|---|
| Age (years; mean ± SD) | 66.5 ± 13.6 | 52.3 ± 15.1 | 72.4 ± 8.5* | 74.3 ± 7.9* | 63.9 ± 10.2* | 67.4 ± 8.0* | 58.5 ± 5.8 |
| Sex (% female) | 1147 (57.9) | 266 (53.7) | 459 (54.8) | 201 (71.0)* | 44 (51.8) | 172 (62.5) | 5 (83.3) |
| BMI (% normal range) | 708 (40.5) | 144 (29.1) | 299 (38.2) | 121 (46.0) | 20 (32.3) | 120 (44.9) | 4 (66.7) |
| Education (years; mean ± SD) | 10.9 ± 5.2 | 15.5 ± 2.9 | 8.3 ± 4.4* | 7.6 ± 4.9* | 12.1 ± 4.6* | 13.2 ± 3.2* | 15.3 ± 2.1 |
| MMSE (mean ± SD) | 24.8 ± 4.3 | 29.4 ± 0.8 | 25.5 ± 3.4* | 20.2 ± 4.4* | 22.1 ± 4.1* | 27.5 ± 3.0* | NA |
| MoCA (mean ± SD) | 23.5 ± 8.8 | 25.1 ± 8.1 | 27.3 ± 2.3 | 14.1 ± 8.0* | 15.1 ± 8.7* | 26.3 ± 2.7 | 27.5 ± 0.7 |
| *APOE* ε4 (% carriers) | 440 (30.0) | 25 (26.3) | 252 (30.3) | 93 (36.6) | 12 (36.4) | 58 (22.9) | NA |
| **Biomarkers (RFU, median ± IQR)** |  |  |  |  |  |  |  |
| SEMA3G | 18131.0 ± 9259.7 | 21071.1 ± 8551.0 | 16318.7 ± 6992.5* | 18108.2 ± 7644.1* | 18231.4 ± 5678.1* | 19713.4 ± 5614.6* | 19096.5 ± 3390.2 |
| NfL | 985.3 ± 563.9 | 730.9 ± 374.7 | 1004.3 ± 474.0* | 1256.4 ± 611.7* | 1714.9 ± 1860.8* | 992.3 ± 480.7* | 4434.4 ± 1441.5* |
| GFAP_1 | 40.8 ± 12.5 | 34.6 ± 9.4 | 40.9 ± 11.7* | 44.7 ± 13.5* | 39.3 ± 12.1 | 44.9 ± 9.3* | 33.9 ± 14.4 |
| GFAP_2 | 1545.6 ± 596.4 | 1239.7 ± 524.2 | 1664.9 ± 550.4* | 1751.5 ± 529.8* | 1783.2 ± 703.1* | 1380.3 ± 477.4* | 1787.4 ± 470.0* |

*Note.* All clinical data are presented as mean ± standard deviation or frequency (%). \* Denotes significance compared to Control. CSF biomarkers are represented as median ± IQR. Abbreviations: AD, Alzheimer's disease; ALS, amyotrophic lateral sclerosis; *APOE*, apolipoprotein E; BMI, body mass index; FTD, frontotemporal dementia; GFAP, glial fibrillary acidic protein (SOMAmers 1 and 2); MCI, mild cognitive impairment; MMSE, Mini-Mental State Examination; MoCA, Montreal Cognitive Assessment; NA, not applicable (no data); NfL, neurofilament light chain; PD, Parkinson's disease; pTau, phosphorylated tau; RFU, relative fluorescence units; SD, standard deviation; SEMA3G, semaphorin 3G. Missing data in the total sample (N = 1982) were as follows: BMI (11.9%); education (0.9%); MMSE (35.8%); MoCA (74.3%); [MMSE + MoCA] (10.1%); *APOE* ε4 carriers (26.0%); GFAP_1 (3.7%). Those with >20% missing data across characteristics and biomarkers in subgroups were: BMI (Control, FTD); MMSE + MoCA (PD, ALS); *APOE* ε4 carrier (Control, AD, FTD, ALS).

**Table 2.** GNPC plasma cohort characteristics with biomarker levels.

|  | **Total sample (N = 6917)** | **Control (n = 1211)** | **MCI (n = 2113)** | **AD (n = 2653)** | **FTD (n = 180)** | **PD (n = 516)** | **ALS (n = 244)** |
|---|---|---|---|---|---|---|---|
| Age (years; mean ± SD) | 71.4 ± 11.0 | 67.2 ± 14.2 | 71.2 ± 9.0* | 75.4 ± 9.0* | 64.0± 10.5* | 70.1 ± 9.4* | 58.3 ± 10.8* |
| Sex (% female) | 3768 (54.5) | 719 (59.4) | 1103 (52.2)* | 1517 (57.2) | 84 (46.7)* | 252 (48.8)* | 93 (38.1)* |
| BMI (% normal range) | 2281 (43.1) | 464 (38.2) | 787 (37.2) | 683 (25.7)* | 43 (23.9) | 230 (48.1)* | 74 (34.7) |
| Education (years; mean ± SD) | 12.6 ± 4.6 | 14.1 ± 3.3 | 11.7 ± 5.0* | 12.2 ± 5.0* | 13.0 ± 4.1* | 14.0 ± 3.6 | 15.6 ± 2.1 |
| MMSE (mean ± SD) | 25.3 ± 5.9 | 28.7 ± 1.5 | 26.7 ± 2.8* | 22.6 ± 7.7* | 22.7 ± 4.2* | 26.6 ± 4.4* | NA |
| MoCA (mean ± SD) | 18.7 ± 12.4 | NA | 24.9 ± 10.2 | 15.1 ± 13.9 | 14.9 ± 11.8 | 25.7 ± 5.0 | 25.3 ± 3.6 |
| *APOE* ε4 (% carriers) | 2235 (44.8) | 270 (40.5) | 680 (32.2) | 1107 (41.7)* | 26 (32.1) | 111 (27.5) | 41 (22.2)* |
| **Biomarkers (RFU, median ± IQR)** |  |  |  |  |  |  |  |
| SEMA3G | 1583.0 ± 364.9 | 1607.6 ± 371.0 | 1585.6 ± 326.8 | 1581.4 ± 411.7* | 1500.0 ± 370.9* | 1567.2 ± 305.1 | 1536.6 ± 375.9* |
| NfL | 508.7 ± 210.2 | 502.3 ± 199.1 | 521.1 ± 199.6 | 484.0 ± 225.1 | 535.1 ± 169.2* | 555.5 ± 193.7* | 556.2 ± 188.0 |
| GFAP_1 | 86.3 ± 28.8 | 85.2 ± 28.1 | 87.8 ± 27.6 | 86.8 ± 30.3 | 82.3 ± 21.4 | 83.9 ± 27.4 | 85.4 ± 25.7 |
| GFAP_2 | 432.4 ± 77.4 | 424.9 ± 57.15 | 429.3 ± 73.6 | 439.6 ± 97.0 | 436.2 ± 72.9 | 423.4 ± 65.2 | 446.6 ± 64.2 |

*Note.* All clinical data are presented as mean ± standard deviation or frequency (%). \* Denotes significance compared to Control. Plasma biomarkers are represented as median ± IQR. Abbreviations: AD, Alzheimer's disease; ALS, amyotrophic lateral sclerosis; *APOE*, apolipoprotein E; BMI, body mass index; FTD, frontotemporal dementia; GFAP, glial fibrillary acidic protein (SOMAmers 1 and 2); MCI, mild cognitive impairment; MMSE, Mini-Mental State Examination; MoCA, Montreal Cognitive Assessment; NA, not applicable (no data); NfL, neurofilament light chain; PD, Parkinson's disease; pTau, phosphorylated tau; RFU, relative fluorescence units; SD, standard deviation; SEMA3G, semaphorin 3G. Missing data in the total sample (N = 6917) were as follows: BMI (25.4%); Education (29.9%); MMSE (14.5%); MoCA (94.0%); [MMSE + MoCA] (8.5%); *APOE* ε4 carrier (27.9%); NfL (2.8%); GFAP_1 (2.8%). Those with >20% missing data across characteristics and biomarkers in subgroups were: BMI (AD, FTD); Education (AD, PD, ALS); MMSE + MoCA (PD, ALS); *APOE* ε4 carrier (Control, MCI, AD, FTD, PD).

In CSF, all groups were significantly older than Control except for ALS, and those with AD were older than all other groups except MCI [F(5, 1976) = 266.4, p < 2.2e-16]. MCI were also older than FTD, PD and ALS. There was a higher proportion of females in AD compared to Control, MCI and FTD (χ^2^(5) = 32.2, p = 5.4e-6). No differences in the proportion of normal BMI were found. All except ALS were significantly less educated than Control, with the AD group being the least educated compared to all other groups except MCI [F(5, 1958) = 257.2, p < 2.2e-16]. Those with MCI were also less educated than FTD, PD and ALS. All pairwise comparisons by group were significant in MMSE scores [F(4, 1267) = 172.5, p < 2.2e-16]. In contrast, AD and FTD groups performed worse on the MoCA compared to Control, MCI and PD [F(5, 503) = 23.5, p < 2.2e-16]. The AD group had a higher proportion of *APOE* ε4 carriers than PD (χ^2^(4) = 12.6, p = 1.3e-2).

The age of each diagnostic group in the plasma cohort was significantly different in all pairwise comparisons [F(5, 6911) = 220.1, p < 2.2e-16], except for MCI versus PD. Those with ALS were the youngest, followed by FTD, Control, PD, MCI and AD. There was a higher proportion of females in Control versus all other groups except AD (χ^2^(5) = 61.3, p = 6.5e-12), as well as in AD compared to MCI, PD and ALS. BMI ranges were different in AD and PD compared to Control, while the FTD group had a lower proportion of normal range BMI compared to AD and PD (χ^2^(15) = 73.2, p = 1.2e-9). Those with MCI or AD were less educated than Control and PD [F(5, 4841) = 50.4, p < 2.2e-16]. The FTD group was also less educated than Control and MCI, and those with AD were less educated than those with MCI. Performance on the MMSE differed in all pairwise comparisons by group [F(4, 5907) = 316.3, p < 2.2e-16], except for MCI and PD, and AD and FTD. In contrast AD and FTD groups performed worse on the MoCA compared to MCI and PD [F(4, 409) = 19.4, p = 1.2e-14]. There were fewer *APOE* ε4 carriers in ALS compared to Control, but more in AD compared to Control, MCI, FTD, PD and ALS, as well as in MCI compared to PD and ALS (χ^2^(5) = 223.6, p < 2.2e-16).

### 3.2 CSF and plasma SEMA3G levels across neurodegenerative conditions

CSF SEMA3G levels were significantly lower in MCI, AD, FTD and PD compared to Control [F(5, 1974) = 43.9, p = 4.6e-43] while adjusting for age and sex, lower in MCI compared to AD and PD, and lower in AD compared to PD (Figure 1, Supplementary Table 1). Similarly, plasma SEMA3G levels were significantly lower in AD, FTD and ALS compared to Control (F(5, 5287) = 7.7, p = 2.9e-7) while adjusting for age, sex and BMI, and lower in AD and ALS compared to MCI (Supplementary Table 2). Hence, SEMA3G was consistently lower in AD and FTD across CSF and plasma. Despite the group-level decrease in plasma SEMA3G in AD compared to Control, we also noted a cluster of AD cases with high SEMA3G that were not observed in other disease groups. An additional analysis was therefore performed categorising cases into those with high (n=131) or low (n=6786) SEMA3G using a relative protein expression threshold of 3200, which separated only the high cluster of AD cases from those with other diagnoses. Univariate t-tests corrected for Bonferroni FDR (Supplementary Table 3) revealed that the high SEMA3G group were younger (t = 5.8, p = 1.2e-8, q = 4.0e-7), less educated (t = 9.7, p = 1.0e-20, q = 3.5e-19), smoked less (t = 25.7, p = 1.5e-134, q = 5.0e-133), and were associated with a lack of the following comorbidities: atrial fibrillation (t = 24.4, p = 1.5e-128, q = 5.0e-127), traumatic brain injury (t = 20.9, p = 2.3e-94, q = 7.7e-93), myocardial infarction (t = 18.9, p = 8.8e-78, q = 3.0e-76), congestive heart failure (t = 18.2, p = 6.1e-73, q = 2.1e-71), transient ischaemic attack (t = 14.2, p = 4.2e-45, q = 1.4e-43), angina (t = 14.1, p = 2.0e-44, q = 6.9e-43), and chronic obstructive pulmonary disease (t = 13.7, p = 2.6e-42, q = 8.8e-41). High plasma SEMA3G participants (>3200 relative protein expression) were only observed at baseline.

**Figure 1.**
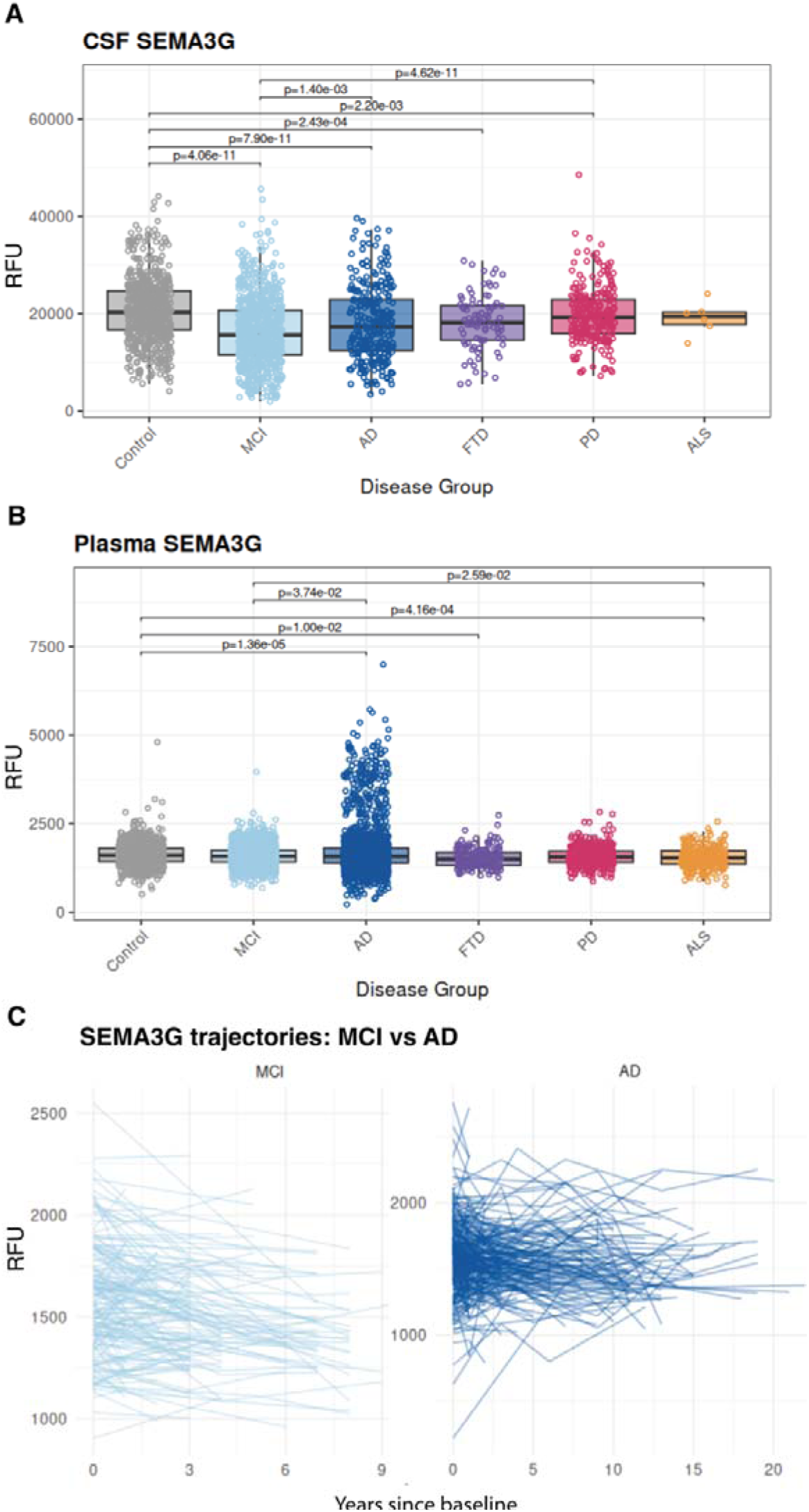
SEMA3G levels are consistently lower in AD and FTD across CSF and plasma, and decrease over time in the plasma of MCI and AD cases. Box plots illustrating protein expression levels of (A) CSF (N = 1982) and (B) plasma (N = 6917) SEMA3G across cognitively unimpaired Control and patients with MCI, AD, FTD, PD and ALS in the Global Neurodegeneration Proteomics Consortium. Significance annotations are shown and derived from one-way ANOVAs adjusting for age, sex and BMI (plasma only) and Tukey’s post hoc (p < 0.05). Box middle line and limits indicate median ± IQRs, respectively, for each group. (C) Individual plasma SEMA3G trajectories over time (years) in MCI and AD. Plots illustrate cases with at least two timepoints, with complete age and sex data. Linear mixed effects models adjusted for age and sex. Abbreviations: AD, Alzheimer’s disease; ALS, amyotrophic lateral sclerosis; ANOVA, analysis of variance; BMI, body mass index; CSF, cerebrospinal fluid; FTD, frontotemporal dementia; IQR, interquartile range; MCI, mild cognitive impairment; PD, Parkinson’s disease; RFU, relative fluorescence units, SEMA3G, semaphorin 3G.

We then examined plasma SEMA3G trajectories over time across the different diagnostic groups. There were 2250 samples in the longitudinal cohort, of which 849 were unique participants. The sample sizes at each timepoint were as follows: t1 (777), t2 (808), t3 (438), t4 (200), t5 (27). The sample sizes for each group were: MCI (391), AD (792), FTD (21), PD (265) and ALS (922). Using linear mixed effects models, we assessed how plasma SEMA3G differed by current diagnosis over time, while adjusting for age and sex, as well as for sample and years since baseline as random effects. SEMA3G declined significantly over time in MCI (β =-25.2, t =-6.9, p = 1.0e-11) and AD (β =-4.4, t =-2.0, p = 4.98e-2) (Supplementary Table 4).

For cross-sectional associations between SEMA3G and covariates in multivariate models, CSF SEMA3G decreased with increasing age [F(1, 1974) = 12.7, p = 3.7e-4]. Plasma SEMA3G decreased with increasing age [F(1, 5287) = 7.8, p = 5.4e-3], was lower in females compared to males [F(1, 5287) = 10.0, p = 1.6e-3], and showed an increasing trend with increasing BMI [F(3, 5287) = 3.2, p = 2.2e-2], as the obese group had higher SEMA3G than the normal (p = 2.5e-2) and underweight (p = 7.7e-3) groups, and the overweight group also had higher levels than the underweight group (p = 4.5e-2). SEMA3G did not correlate with NfL, GFAP or amyloid precursor protein (APP) in either CSF or plasma or within subgroups. Using a threshold of partial R^2^ > 0.25 (q < 0.05), there were 11 plasma proteins in Control that were significantly but weakly associated with SEMA3G, whereas there were two proteins (Kringle-containing transmembrane protein 1 [KREMEN1, partial R^2^ = 0.27, q = 8.5e-35] and Plexin D1 [PLXND1, partial R^2^ = 0.25, q = 3.7e-31]) weakly associated with SEMA3G in AD (Supplementary Table 5). As CAA has been shown to be associated with *APOE* ε2 and ε4 alleles [17], we examined if there were any allele group differences in SEMA3G in CSF and plasma. No significant differences were found in CSF, while those with ε3ε4 had higher plasma SEMA3G than those with ε3ε3 (Kruskal-Wallis χ^2^(6) = 14.7, p = 0.02, q = 0.05).

When assessing relationships to comorbidities while adjusting for covariates, CSF SEMA3G was significantly increased in those with hypertension compared to those without after correcting for FDR [t(1274) = 7.4, p = 2.0e-13, q = 6.9e-11] (Supplementary Table 6). In contrast, plasma SEMA3G was significantly lower in individuals with hypertension [t(7331) = 8.0, p = 2.0e-15, q = 1.1e-12] and hyperlipidaemia [t(6609) = 7.8, p = 5.6e-15, q = 3.2e-12].

### 3.3 Healthy Brain Ageing cohort

As reported in Table 3, the HBA cohort (n = 161) comprised predominantly cognitively normal (mean MMSE score = 28.1), well-educated (mean education = 14.8) older adults approximately 70 years old, with approximately one-third carrying at least one *APOE* ε4 allele. Relevant to this sample of community-dwelling older adults attending a memory clinic, mean plasma pTau217 concentrations (0.6 pg/mL) indicated an intermediate burden of AD pathology according to the criteria proposed by Ashton et al. (2024). Whole brain neuroimaging indicated a burden of cerebrovascular disease, reflected by a mean of 53.5 (SD = 41.5) white matter hyperintensity lesions per participant.

**Table 3:**
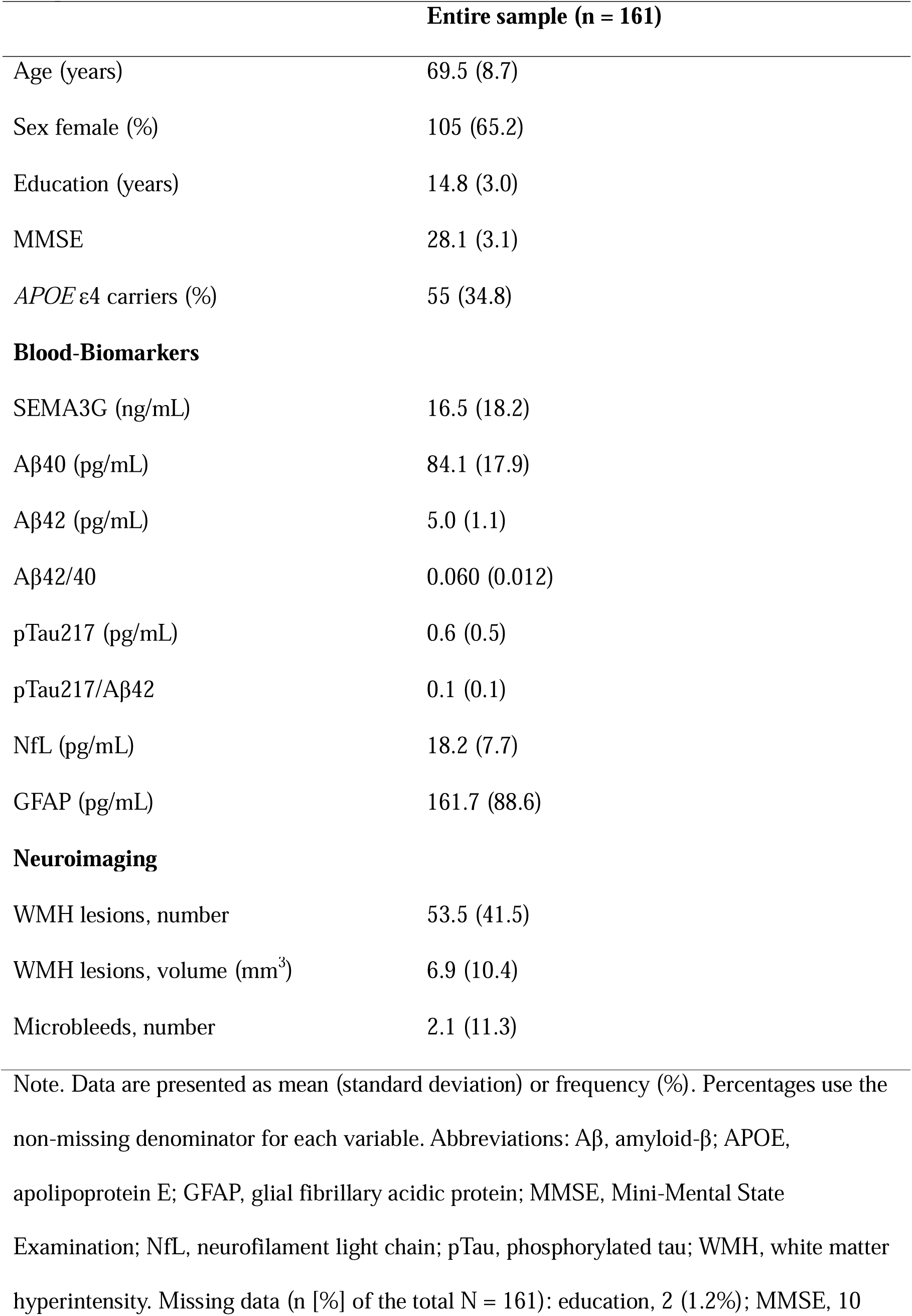
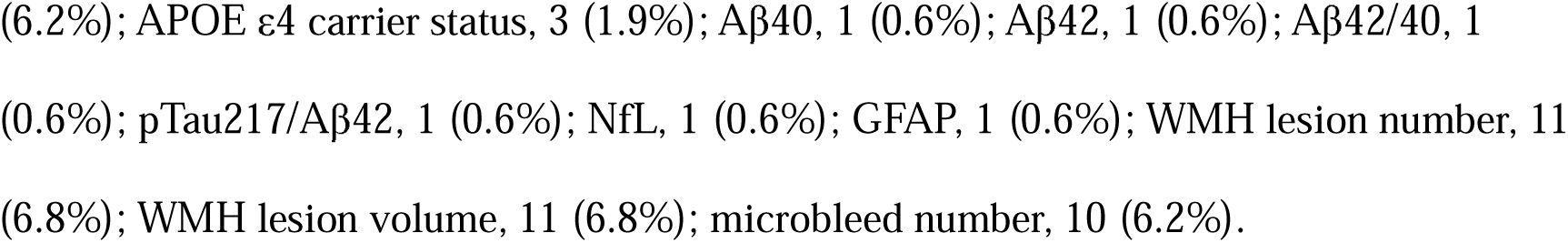
Clinical, blood biomarker, and neuroimaging characteristics of the entire sample.

|  | **Entire sample (n = 161)** |
|---|---|
| Age (years) | 69.5 (8.7) |
| Sex female (%) | 105 (65.2) |
| Education (years) | 14.8 (3.0) |
| MMSE | 28.1 (3.1) |
| *APOE* ε4 carriers (%) | 55 (34.8) |
| **Blood-Biomarkers** |  |
| SEMA3G (ng/mL) | 16.5 (18.2) |
| Aβ40 (pg/mL) | 84.1 (17.9) |
| Aβ42 (pg/mL) | 5.0 (1.1) |
| Aβ42/40 | 0.060 (0.012) |
| pTau217 (pg/mL) | 0.6 (0.5) |
| pTau217/Aβ42 | 0.1 (0.1) |
| NfL (pg/mL) | 18.2 (7.7) |
| GFAP (pg/mL) | 161.7 (88.6) |
| **Neuroimaging** |  |
| WMH lesions, number | 53.5 (41.5) |
| WMH lesions, volume (mm<sup>3</sup>) | 6.9 (10.4) |
| Microbleeds, number | 2.1 (11.3) |

Note. Data are presented as mean (standard deviation) or frequency (%). Percentages use the non-missing denominator for each variable. Abbreviations: Aβ, amyloid-β; APOE, apolipoprotein E; GFAP, glial fibrillary acidic protein; MMSE, Mini-Mental State Examination; NfL, neurofilament light chain; pTau, phosphorylated tau; WMH, white matter hyperintensity. Missing data (n [%] of the total N = 161): education, 2 (1.2%); MMSE, 10 (6.2%); APOE ε4 carrier status, 3 (1.9%); Aβ40, 1 (0.6%); Aβ42, 1 (0.6%); Aβ42/40, 1 (0.6%); pTau217/Aβ42, 1 (0.6%); NfL, 1 (0.6%); GFAP, 1 (0.6%); WMH lesion number, 11 (6.8%); WMH lesion volume, 11 (6.8%); microbleed number, 10 (6.2%).

Table 4 presents the clinical characteristics, plasma biomarker concentrations and neuroimaging measures of the sample stratified by the likelihood of CAA, as indexed by the presence and number of lobar cerebral microbleeds. Age differed significantly across groups, with participants with no MRI evidence of CAA (0 lobar cerebral microbleeds) being younger than those with increasing CAA-associated evidence on MRI. There were no significant differences in plasma SEMA3G concentrations across either the microbleed groups or pTau217 classification groups (negative, indeterminate and positive), and as illustrated in Figure 2. WMH lesion number and volumes were significantly higher in the group with higher CAA-associated evidence on MRI and elevated ARIA risk (≥3 microbleeds) compared to the group with no MRI evidence of CAA (0 microbleeds).

**Figure 2.**
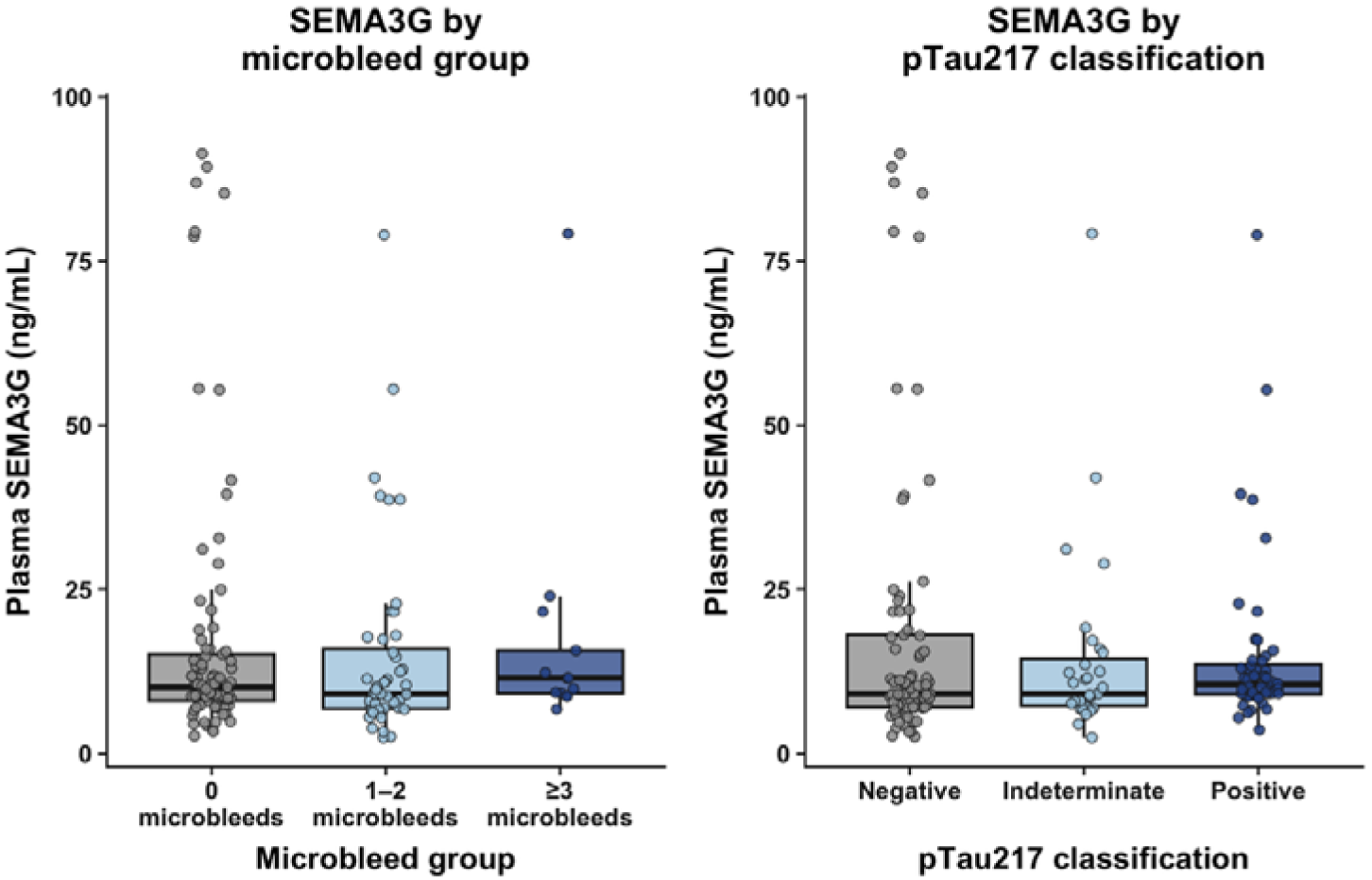
Plasma SEMA3G concentrations (ng/mL) stratified by cerebral microbleed burden and plasma pTau217 classification. Cerebral microbleed burden (left) was categorised as 0 (no CAA-associated evidence on MRI), 1-2 (low CAA-associated evidence on MRI) or ≥3 microbleeds (higher CAA-associated evidence on MRI and elevated ARIA risk). Plasma pTau217 status (right) was classified as negative, indeterminate or positive according to the Ashton et al. (2024) criteria. Boxes indicate the interquartile range, with horizontal lines representing the median. Whiskers extend to the most extreme values within 1.5 times the interquartile range of the box limits. Overall group comparisons used Kruskal-Wallis tests, and pairwise comparisons used Dunn tests with Holm adjustment. No statistically significant pairwise differences were observed in either panel (all Holm-adjusted p > 0.05).

**Table 4.** Clinical, blood biomarker, and neuroimaging characteristics stratified by CAA-associated evidence on MRI.

| Characteristic | **Total Subsample with microbleed data**<br>**N = 150<sup>1</sup>** | **A**<br>**0 lobar cerebral microbleeds**<br>**(No CAA-associated evidence on MRI)**<br>**N = 89<sup>1</sup>** | **B**<br>**1-2 lobar cerebral microbleeds**<br>**(Low CAA-associated evidence on MRI)**<br>**N = 48<sup>1</sup>** | **C**<br>**≥ 3 lobar cerebral microbleeds**<br>**(higher CAA-associated evidence on MRI and elevated ARIA risk)**<br>**N = 13<sup>1</sup>** | **Statistic<sup>†</sup>** | **P value<sup>†</sup>** | **Pairwise comparison** |
|---|---|---|---|---|---|---|---|
| Age (years) | 69.5 (8.7) | 67.3 (8.8) | 72.0 (7.7) | 75.2 (6.7) | 14.2 | <0.001 | A<B, A<C |
| Sex, female | 100 (66.7%) | 64 (71.9%) | 31 (64.6%) | 5 (38.5%) | 5.8 | 0.054 |  |
| Education (years) | 14.6 (3.1) | 14.4 (3.0) | 14.8 (3.2) | 15.1 (3.5) | 1.4 | 0.504 |  |
| MMSE | 28.2 (2.7) | 28.0 (3.2) | 28.6 (1.8) | 28.2 (2.3) | 0.2 | 0.903 |  |
| APOE ε4 carriers | 51 (34.7%) | 30 (34.5%) | 15 (31.9%) | 6 (46.2%) | 0.9 | 0.633 |  |
| **Blood-Biomarkers** |  |  |  |  |  |  |  |
| SEMA3G (ng/mL) | 16.7 (18.8) | 17.7 (20.7) | 14.8 (14.8) | 17.6 (19.2) | 2.2 | 0.340 |  |
| Aβ40 (pg/mL) | 84.6 (17.6) | 81.8 (15.1) | 86.6 (17.1) | 95.9 (28.5) | 4.6 | 0.102 |  |
| Aβ42 (pg/mL) | 5.0 (1.1) | 5.1 (1.1) | 4.9 (1.1) | 5.1 (1.4) | 0.2 | 0.909 |  |
| Aβ42/40 | 0.060 (0.012) | 0.063 (0.012) | 0.057 (0.012) | 0.055 (0.012) | 6.6 | 0.036 |  |
| pTau217 (pg/mL) | 0.6 (0.5) | 0.5 (0.4) | 0.6 (0.6) | 0.7 (0.4) | 4.6 | 0.100 |  |
| pTau217/Aβ42 | 0.1 (0.1) | 0.1 (0.1) | 0.1 (0.2) | 0.1 (0.1) | 4.7 | 0.097 |  |
| NfL (pg/mL) | 18.2 (7.7) | 17.3 (7.8) | 18.9 (6.7) | 21.7 (9.4) | 4.8 | 0.090 |  |
| GFAP (pg/mL) | 163.0 (89.0) | 150.3 (80.2) | 184.9 (105.5) | 168.6 (64.6) | 3.7 | 0.154 |  |
| **Neuroimaging** |  |  |  |  |  |  |  |
| WMH lesions, number | 52.7 (41.7) | 46.9 (40.0) | 55.3 (40.2) | 83.9 (46.0) | 9.2 | 0.010 | A<C |
| WMH lesions, volume (mm<sup>3</sup>) | 6.9 (10.6) | 5.4 (8.6) | 8.1 (13.0) | 12.9 (12.0) | 11.1 | 0.004 | A<C |
| Microbleeds, number | 2.1 (11.3) | 0.0 (0.0) | 1.4 (0.5) | 18.8 (35.4) | 144.3 | <0.001 | A<B, A<C, B<C |

*Note.* Data are presented as mean (standard deviation) or frequency (%)
Abbreviations: Aβ, amyloid β; APOE, apolipoprotein E; ARIA, amyloid-related imaging abnormalities; CAA, cerebral amyloid angiopathy; GFAP, glial fibrillary acidic protein; MMSE, Mini-Mental State Examination; NfL, neurofilament light chain; pTau, phosphorylated tau; WMH, white-matter hyperintensity.
<sup>1</sup>Mean (SD); n (%)
<sup>†</sup>Anova with post hoc t-tests, Kruskal–Wallis with post hoc Dunn tests, or Chi-Squared tests, as appropriate.

As illustrated in Figure 3, plasma SEMA3G was not significantly correlated with any CAA-associated neuroimaging measure on MRI (lobar microbleed count, WMH lesion count or volume) or plasma biomarker (Aβ40, Aβ42, Aβ42/40, pTau217, pTau217/Aβ42, NfL or GFAP). In contrast, pTau217, pTau217/Aβ42, NfL and GFAP were strongly positively correlated with WMH lesion count and volume (Spearman’s ρ =.61-.73, all *p* <.001), while Aβ40, pTau217/Aβ42, NfL and GFAP showed weaker positive correlations with microbleed count (Spearman’s ρ =.18-.21, all *p* <.05). When stratifying groups based on CAA-associated evidence on MRI (Figure 4), plasma biomarker-neuroimaging associations revealed in Figure 3 were most evident in the group with no CAA-associated evidence on MRI (0 microbleeds) and increased risk (1-2 microbleeds) groups, while associations were less consistent in the higher risk (≥ 3 microbleeds) group. Notably, SEMA3G was not associated with microbleed count, WMH lesion count or volume in any group. SEMA3G was also unrelated to the other plasma biomarkers when stratifying groups based on CAA-associated evidence on MRI, but was inversely correlated with NfL in the higher risk (≥ 3 microbleeds) group (Spearman’s ρ =-.71, *p* <.01), although this finding should be interpreted cautiously given the small group size (n = 13).

**Figure 3:**
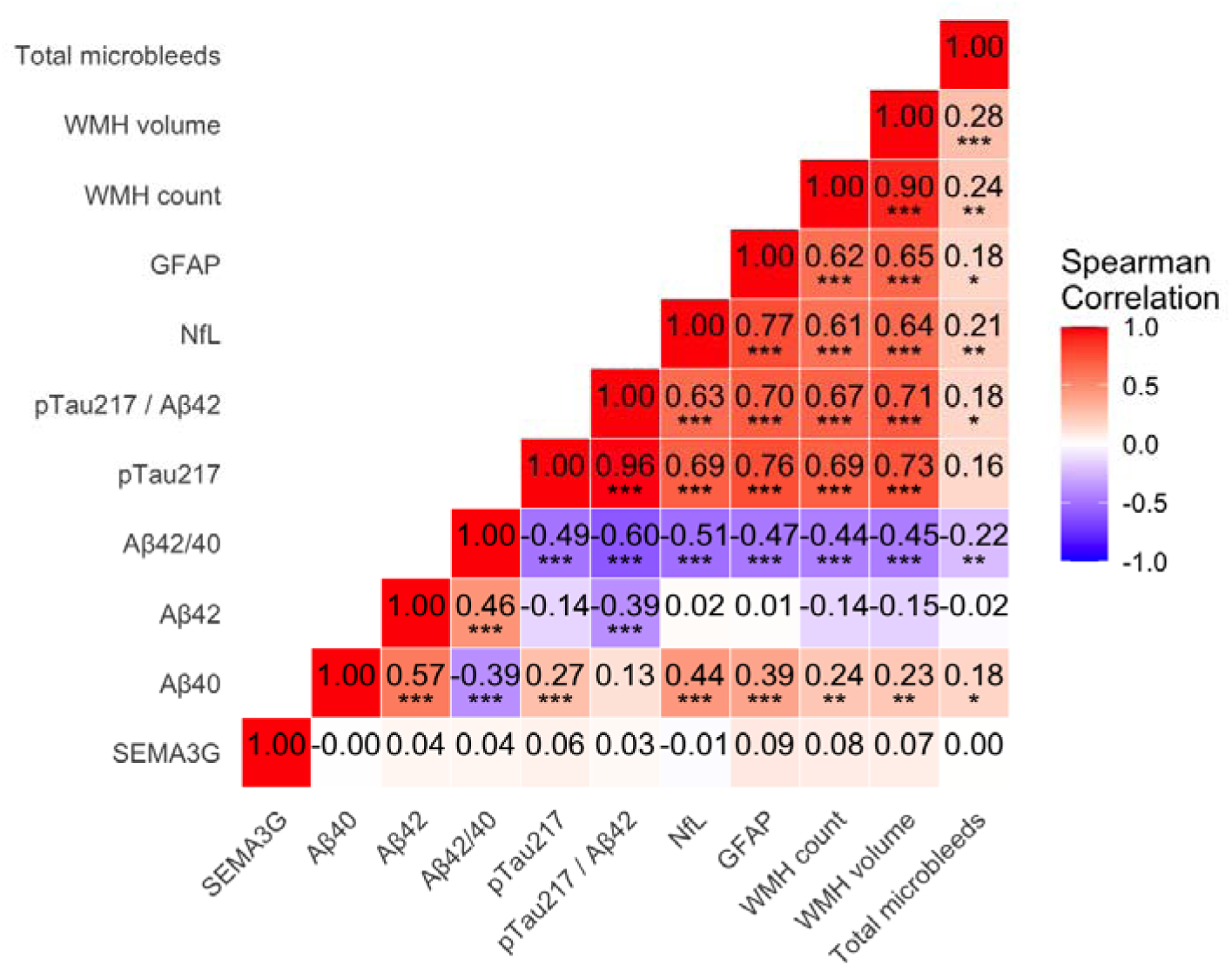
Spearman correlation matrix between plasma biomarkers (including SEMA3G) and neuroimaging markers of cerebral amyloid angiopathy in all samples with microbleed data (n = 150). Correlations were calculated using Spearman’s rank correlation in participants with available microbleed data (n = 150). Neuroimaging markers include total microbleed count, white matter hyperintensity (WMH) count, and WMH volume. p values are denoted as *p* < 0.05*, *p* < 0.01**, and *p* < 0.001***. Abbreviations: Aβ, amyloid β; CAA, cerebral amyloid angiopathy; GFAP, glial fibrillary acidic protein; MMSE, Mini-Mental State Examination; NfL, neurofilament light chain; pTau, phosphorylated tau; WMH, white-matter hyperintensity.

**Figure 4:**
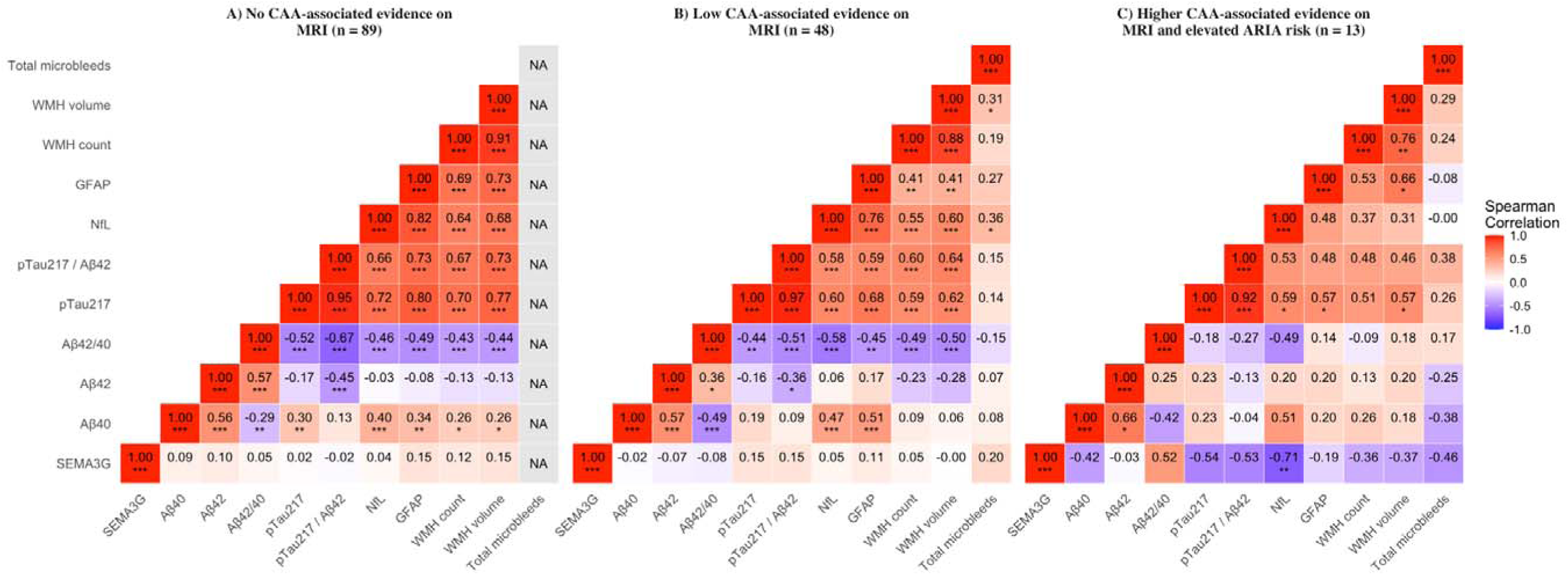
Spearman correlation matrices between plasma biomarkers (including SEMA3G) and neuroimaging markers of cerebral amyloid angiopathy stratified by CAA-associated evidence on MRI. Correlations were calculated using Spearman’s rank correlation in participants with available microbleed data. Neuroimaging markers include total microbleed count, white matter hyperintensity (WMH) count, and WMH volume. p values are denoted as *p* < 0.05*, *p* < 0.01**, and *p* < 0.001***. Abbreviations: Aβ, amyloid β; ARIA, amyloid-related imaging abnormalities; CAA, cerebral amyloid angiopathy; GFAP, glial fibrillary acidic protein; MMSE, Mini-Mental State Examination; NA, not estimable; NfL, neurofilament light chain; pTau, phosphorylated tau; WMH, white-matter hyperintensity.

## 4. Discussion

This is the first study to investigate SEMA3G as a viable central or peripheral biofluid biomarker of CAA across neurodegenerative conditions, and examined plasma SEMA3G in relation to MRI features associated with CAA in a memory clinic cohort. While we identified significantly lower CSF and plasma SEMA3G levels in AD and FTD cases in the larger GNPC cohort, as well as significant longitudinal declines in both MCI and AD, plasma SEMA3G levels in the HBA memory clinic cohort did not correlate with CAA-associated markers on MRI, including lobar cerebral microbleeds and white matter hyperintensities, or with plasma Aβ40 or Aβ42. Therefore, in these cohorts, our findings do not convincingly support plasma SEMA3G as a peripheral biomarker for identifying individuals with higher sporadic CAA-associated burden or elevated ARIA risk in a memory clinic setting.

Here, we report for the first time that SEMA3G concentrations are significantly lower in both the CSF and plasma of older adults living with AD, lower in the CSF of those living with MCI, and decline significantly over time in both MCI and AD. Indeed, our observation that SEMA3G was significantly decreased in the CSF of older adults with AD in the larger GNPC cohort is consistent with a previous small study [18]. While our findings reveal that plasma SEMA3G levels significantly differ between neurodegenerative conditions, including across the AD continuum, the substantial overlap may suggest limited disease specificity. That is, autopsy studies have reported CAA in approximately 80% of AD cases, but it is also found in individuals with dementia with Lewy Bodies (∼50-70%), PD (∼24-45%), and MCI (∼23%) [4, 19, 20]. Given this substantial overlap, it may be difficult to establish whether any sporadic CAA-related changes in biofluid SEMA3G concentrations are specific to AD or another particular neurodegenerative condition.

The absence of an association between plasma SEMA3G and MRI markers of probable CAA was unexpected given our previous findings showing a strong correlation between SEMA3G and CAA severity in the brain [10, 11]. That is, our previous neuropathological findings revealed a marked increase of SEMA3G in all CAA-affected blood vessels, even those with mild CAA, suggesting that elevated SEMA3G is a highly specific marker of both early and advanced CAA [10]. There are two potential explanations for our discrepancy in findings between brain tissue and biofluids. First, CAA-associated SEMA3G accumulation may be localised to CNS vessel walls [21] without corresponding increases in plasma or CSF, particularly in cases of sporadic CAA that develop slowly with age. A conceptual parallel is provided by Aβ42, for which sequestration in cerebral amyloid deposits is thought to contribute to reduced biofluid concentrations. Although a similar mechanism has not been established for SEMA3G, the numerically lower plasma SEMA3G concentrations observed among participants with greater lobar cerebral microbleed burden in the HBA memory clinic cohort are compatible with this hypothesis. Second, studies in humans and mice have shown that SEMA3G is expressed beyond the CNS, including by endothelial cells across multiple organs, as well as by adipocytes, glomerular podocytes, pancreatic β cells and immune cells [22–25]. SEMA3G derived from these peripheral sources may obscure changes in plasma concentrations arising specifically from CAA-affected CNS vessels, potentially contributing to the discrepancy between brain tissue and plasma findings.

Our finding that lower plasma Aβ42/40 and WMHs is associated with lobar cerebral microbleeds, corroborates previous reports [26–28], and is consistent with overlapping AD-related and cerebrovascular pathology. Moreover, we report for the first time an association between lower plasma SEMA3G and higher NfL concentrations in individuals with greater MRI evidence of sporadic CAA. While we interpret this finding cautiously given the small subgroup size, the association is biologically plausible. That is, studies in mice have shown that SEMA3G supports communication between endothelial cells and neurons and helps maintain synaptic function. SEMA3G also signals through NRP2/PLXND1 receptors to influence endothelial behaviour and vascular remodelling [29]. Given that higher NfL concentrations reflect axonal injury, our observation suggests a possible link between lower plasma SEMA3G and greater neuronal injury in this group. Indeed, our GNPC findings provide further context for this relationship. SEMA3G clustered with other semaphorins, including SEMA6A, SEMA6B and SEMA7A, which may reflect shared roles in neuronal, vascular and immune processes [30]. SEMA3G was also associated with proteins involved in neuronal signalling, including neurotensin receptor 1, tropomyosin receptor kinase B, neurotrimin and immunoglobulin superfamily member 3 [31–34]. These associations are consistent with the involvement of SEMA3G in neuronal function. However, whether lower plasma SEMA3G reflects reduced neuronal support, changes in response to injury or another shared process remains to be studied.

Our study has several limitations that warrant consideration. First, in the larger GNPC cohort CAA diagnostic information was absent, leaving only *APOE* ε4 carrier status and cardiovascular conditions such as stroke and hypertension, as the closest proxy correlates of CAA. While *APOE* ε4 carrier status is associated with CAA, hypertension is only a major risk factor for deep cerebral microbleeds, while the association to stroke depends on the anatomical subtype (ischaemic and lobar intracerebral haemorrhages) [35]. Second, the Boston criteria v2.0 are limited by poor sensitivity, especially in early stage patients and those without haemorrhagic lesions [36]. An estimated likelihood of CAA was determined in the HBA cohort based on these criteria, using only the number of lobar cerebral microbleeds. As such, other MRI markers such as superficial siderosis, severe centrum semiovale perivascular spaces, and specific multispot WMH pattern need to be considered [9]. Third, the cohorts used in this study are younger than those typically examined at autopsy, so the prevalence of expected CAA is likely underestimated in our cohorts. In addition, the HBA memory clinic cohort had limited sample sizes, particularly for the group with higher sporadic CAA-associated burden, for which there were only 13 samples.

While our findings do not support plasma SEMA3G as a peripheral biomarker for identifying individuals with higher sporadic CAA-associated burden or elevated ARIA risk in our selected cohorts, future directions can be considered. Future studies should seek to examine subpopulations where CAA can be definitively confirmed. Although hereditary forms of CAA are rare, Dutch-type CAA provides a well-characterised population in which to investigate potential biomarkers. It is caused by an inherited APP E693Q mutation, corresponding to a glutamate-to-glutamine substitution at position 22 of Aβ (E22Q), which promotes vascular amyloid accumulation [37]. Its clinical features overlap with sporadic CAA, including intracerebral haemorrhage and cognitive impairment, but generally develop at a younger age with a more aggressive progression [38]. Because mutation carriers can be identified before symptom onset, these cohorts provide an opportunity to examine plasma SEMA3G across presymptomatic and symptomatic stages and assess its relationship with CAA-associated disease burden. However, any association identified in hereditary CAA would require independent validation in sporadic CAA, as mutation-related differences in Aβ aggregation and clearance may influence biomarker concentrations and their relationship with pathology. Finally, given SEMA3G’s enrichment and high specificity in CAA-positive vessels, future studies should explore its potential as a target for PET imaging of CAA.

In conclusion, while our previous neuropathological findings identified SEMA3G as a highly specific marker of CAA in the brain, the present findings do not support plasma SEMA3G as a biomarker for identifying individuals with greater MRI evidence of sporadic CAA. However, lower CSF and plasma SEMA3G concentrations in AD, lower CSF concentrations in MCI, longitudinal declines in MCI and AD, and associations with other semaphorins and neuronal signalling proteins warrant further investigation. The association between lower plasma SEMA3G and higher NfL in the subgroup with greater lobar cerebral microbleed burden also suggests a possible link with neuronal injury, although this remains exploratory. Future studies should examine these relationships in larger longitudinal cohorts characterised by neuropathology or comprehensive MRI assessment, alongside genetically defined hereditary CAA cohorts.

## Data Availability

The de-identified data that support the findings of this study are available from the corresponding author upon reasonable request, subject to appropriate data-sharing agreements and compliance with the conditions of the study's ethical approvals.

## Acknowledgements

The authors would like to thank all participants of the Healthy Brain Ageing Program. JCM is supported by the Kennards Hire Foundation Dementia Research Fellowship. LS is supported by internal funding from the Brain and Mind Centre, The University of Sydney.

## Consent statement

All research activities were approved by the University of Sydney Human Research Ethics Committee (Protocol numbers: 2012/1873 and 2019/HE000271). All participants provided informed written consent prior to study participation. The study was performed in accordance with the ethical standards as laid down in the 1964 Declaration of Helsinki and its later amendments or comparable ethical standards.

